# Characterizing super-spreading events and age-specific infectiousness of SARS-CoV-2 transmission in Georgia, USA

**DOI:** 10.1101/2020.06.20.20130476

**Authors:** Max SY Lau, Bryan Grenfell, Michael Thomas, Michael Bryan, Kristin Nelson, Ben Lopman

## Abstract

It is imperative to advance our understanding of heterogeneities in the transmission of SARS-CoV-2 such as age-specific infectiousness and super-spreading. To this end, it is important to exploit multiple data streams that are becoming abundantly available during the pandemic. In this paper, we formulate an individual-level spatio-temporal mechanistic framework to integrate individual surveillance data with geo-location data and aggregate mobility data, enabling a more granular understanding of the transmission dynamics of SARS-CoV-2. We analyze reported cases, between March and early May 2020, in five (urban and rural) counties in the State of Georgia USA. First, our results show that the reproductive number reduced to below 1 in about two weeks after the shelter-in-place order. Super-spreading appears to be widespread across space and time, and it may have a particularly important role in driving the outbreak in rural areas and an increasing importance towards later stages of outbreaks in both urban and rural settings. Overall, about 2% of cases were directly responsible for 20% of all infections. We estimate that the infected non-elderly cases (<60) may be 2.78 [2.10, 4.22] times more infectious than the elderly, and the former tend to be the main driver of super-spreading. Our results improve our understanding of the natural history and transmission dynamics of SARS-CoV-2. More importantly, we reveal the roles of age-specific infectiousness and characterize systematic variations and associated risk factors of super-spreading. These have important implications for the planning of relaxing social distancing and, more generally, designing optimal control measures.

**Significance Statement:** There is still considerable scope for advancing our understanding of the epidemiology and ecology of COVID-19. In particular, much is unknown about individual-level transmission heterogeneities such as super-spreading and age-specific infectiousness. We statistically synthesize multiple valuable datastreams, including surveillance data and mobility data, that are available during the current COVID-19 pandemic. We show that age is an important factor in the transmission of the virus. Super-spreading is ubiquitous over space and time, and has particular importance in rural areas and later stages of an outbreak. Our results improve our understanding of the natural history the virus and have important implications for designing optimal control measures.

---

The current COVID-19 pandemic continues to spread and impact countries across the globe. There is still much scope for mapping out the whole spectrum of the epidemiology and ecology of this novel virus. In particular, understanding of heterogeneities in transmission, which is essential for devising effective targeted control measures, is still limited. For instance, much is unknown about the variation of infectiousness among different age groups [1–3]. Also, while super-spreading events have been documented, its impact and variation over space and time and associated risk factors have not yet been systematically characterized [1, 4–6].

For this reason, it is crucial to exploit the growing availability of multiple data streams during the pandemic, from which we may obtain a more comprehensive picture of the transmission dynamics of SARS-CoV-2. For example, shelter-in-place order likely change the movement patterns of a population, by reducing distance of travel. Such a change needs to be taken into account as movement is a key factor that shapes transmission [7]. Failing to capture this change would also bias the estimates of key model parameters including intervention efficacy and transmissibility parameters that are correlated with movement [8–10]. De-identified mobility data from mobile phone users have been made available to state governments and research institutes through partnerships with private companies such as Facebook. Integration of such mobility data with surveillance data would allow us to account for the change in movement, and therefore more accurately infer the transmission dynamics [7, 8, 10, 11]. Geo-spatial location data and detailed spatial distribution of population are also important for capturing heterogeneous mixing in space [8–10]. A key step is to enable individual-level model inference that can properly synthesize these datastreams, which would go beyond most efforts so far that have focused on aggregated level dynamics [2, 11, 12].

In this paper, building on a previous framework we developed for modelling Ebola outbreaks in Western Africa [8, 9], we formulate an individual-level space-time stochastic model that describes the transmission of SARS-CoV-2 and captures the impact of state-wide social distancing measure in the state of Georgia, USA. Our model mechanistically integrates detailed individual-level surveillance data, geo-spatial location data and highly-resolved population density (grid-)data, and aggregate mobility data (see *Study Data)*. We estimate model parameters and unobserved model quantities including infection times and transmission paths using Bayesian data-augmentation techniques in the framework of Markov Chain Monte Carlo (MCMC) (see *Materials and Methods*). Our individual-level modelling framework also allows us to compute population-level epidemiological parameter such as the basic reproductive number *R*_0_ and quantify the degree of super-spreading over space and time.

## Study Data

We analyzed a rich set of COVID-19 surveillance data collected by the Georgia Department of Public Health, between March 1, 2020, and May 3, 2020, in five counties which had the largest numbers of cases. These counties include four (Cobb, DeKalb, Gwinnet, and Fulton) in the metro Atlanta area and one (Dougherty) in rural southwest Georgia. This dataset contains demographic information of 9,559 symptomatic cases which include age, sex and race, and symptom onset times. It also contains geo-location of the residence of cases. The GDPH Institutional Review Board has determined that this analysis is exempt from the requirement for IRB review and approval. Highly-resolved population-density data over 100 × 100meter grids are obtained from *http://www.worldpop.org.uk*., and are used to modulate the spatial spread of the virus (see *Materials and Methods)*. Aggregate mobility data are used to characterize the average change of movement distance within a county before and after the implementation of state-wide social distancing measure. Specifically, we used high-volume mobility data accessed through Facebook’s Data for Good program [13]. These data represent Facebook users in Georgia who have location services enabled on their mobile device. These data provide information on the number of ‘trips’ (and trip distance) that occurred daily among users. A ‘trip’ is defined as a directional vector starting at the location where an individual spent most of their time during the previous 8-hour period and ending at the location where the same individual spent most of their time during the current 8-hour period.

## Results

### Natural History Parameters and Effectiveness of Social Distancing

We estimate that the median value of *R*_0_ across five counties was overall between 3.30 with 95% C.I. [2.34, 5.2] before the shelter-in-place order. Dougherty county in the rural area had the largest prior-intervention *R*_0_ (5.19 [5.01, 5.31]) and time-varying effective reproductive number *R_eff_* at the earlier stage in March 2020 (Figure 1). Our results suggest the shelter-in-place order was effective, and in all the counties the *R_eff_* declined below 1 in about two to three weeks since the intervention. This is consistent with a recent study which shows that effect of changes of mobility on reducing transmission may become noticeable after 9-12 days [14]. It is also worth noting that *R_eff_* appears to begin decline one to two weeks before shelter-in-place order in urban areas and earlier in Dougherty. We also estimate that the incubation period (i.e. waiting time from infection to symptoms onset) has a median value 6.94 days [5.30, 7.37]. These estimates are largely consistent with the literature [15, 16].

**Fig. 1.**
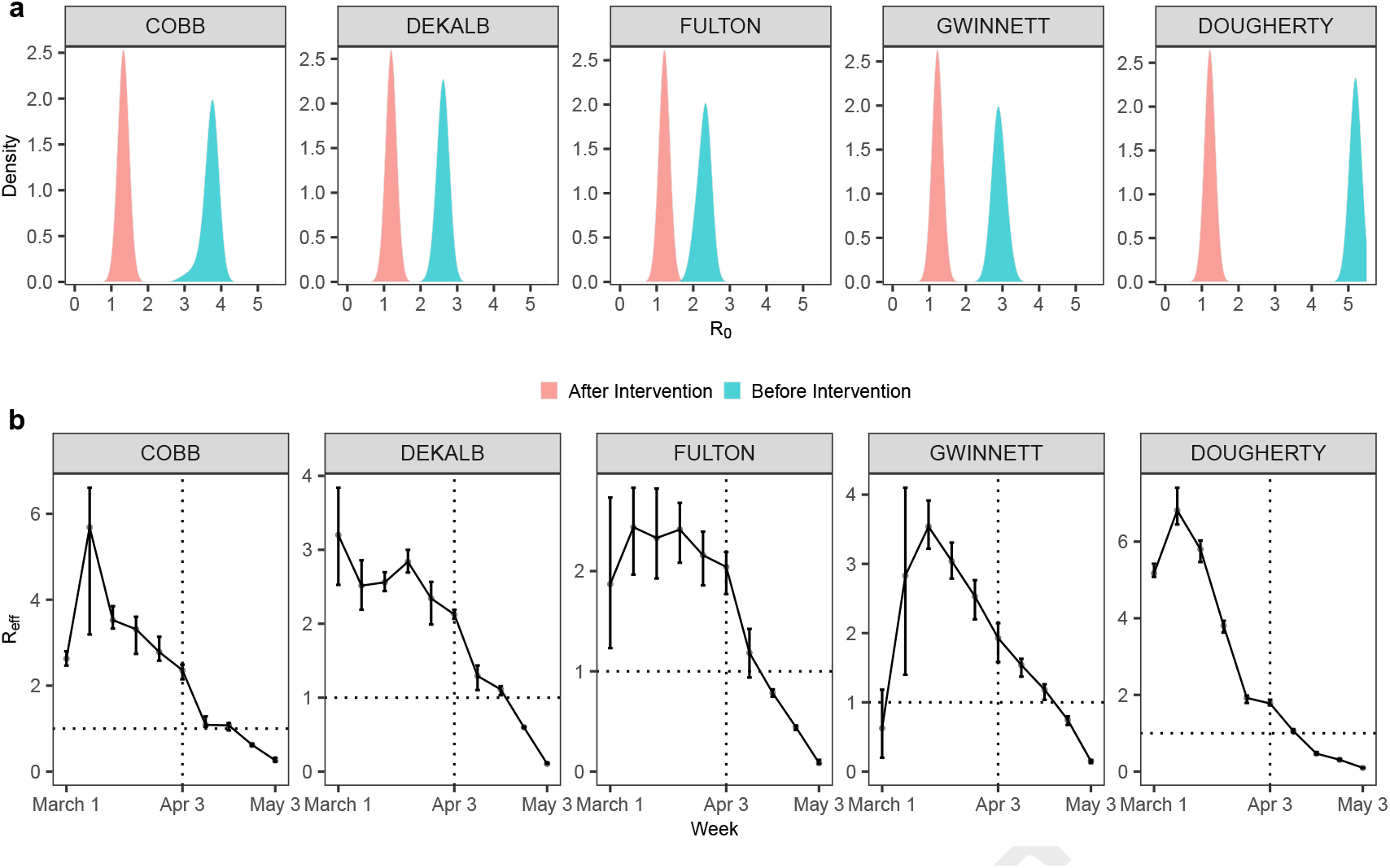
Posterior distribution of basic reproductive number *R*_0_ and the effective reproductive number *R_eff_*, before and after the implementation of state-wide shelter-in-place order on April 3, 2020. Errorbars represent 95% credible interval.

### Systematic Characterization of Super-spreading

Super-spreading refers a phenomenon where certain individuals disproportionately infect a large number of secondary cases relative to an “average” infectious individual (whose infectiousness may be well-represented by *R*_0_). This phenomenon plays a key role in driving the spread of many pathogens including MERS and Ebola [8, 17]. A common measure of the degree of super-spreading is the dispersion parameter *k*, assuming that the distribution of the *offspring* (i.e. number of secondary cases generated) is negative binomial with variance *σ*^2^ = *μ* (1+ *μ/k)* where *μ* is the mean [17]. Generally speaking, a lower *k* corresponds to a higher degree of super-spreading; and *k* less than 1 implies substantial super-spreading. Our framework infers the transmission paths among all cases and therefore naturally generates the offspring distribution of each case (see *Materials and Methods*).

While super-spreading of COVID-19 has been observed [1, 4–6], systematic characterization of its impact and variation (e.g., over time and space) and associated risk factors is lacking. Our results (Figure 2a) suggest that super-spreading is a ubiquitous feature during different periods (before and after the shelter-in-place order) of the outbreak. Super-spreading may have a major impact for the rural area (Dougherty) among all counties (i.e. overall *k* = 0.27 for Dougherty which is the lowest among all counties). Dougherty county has a disproportionately large outbreak compared to other more populated counties – having about only 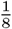 of the population of Cobb county (about 760,000), it has a comparable number of reported cases (1,628). Such an anomaly may be a consequence of the significant super-spreading and large (prior-intervention) *R*_0_ in Dougherty (Figure 1a). This is also consistent with the evidence of super-spreading events due to a funeral in the area [18]. The increasing significance of super-spreading over time also highlights the importance of maintaining social distancing measures that may curtail close contacts (e.g. gatherings with densely packed crowds). Overall, the top 2% of cases (that generate highest number of infections) are responsible directly for about 20% of the total infections. Our results also show that the younger infectees (<60) tends to the main driver of super-spreading (Figure 2b), infectiousness in this age group is also higher (see *Age-specific Infectiousness*).

**Fig. 2.**
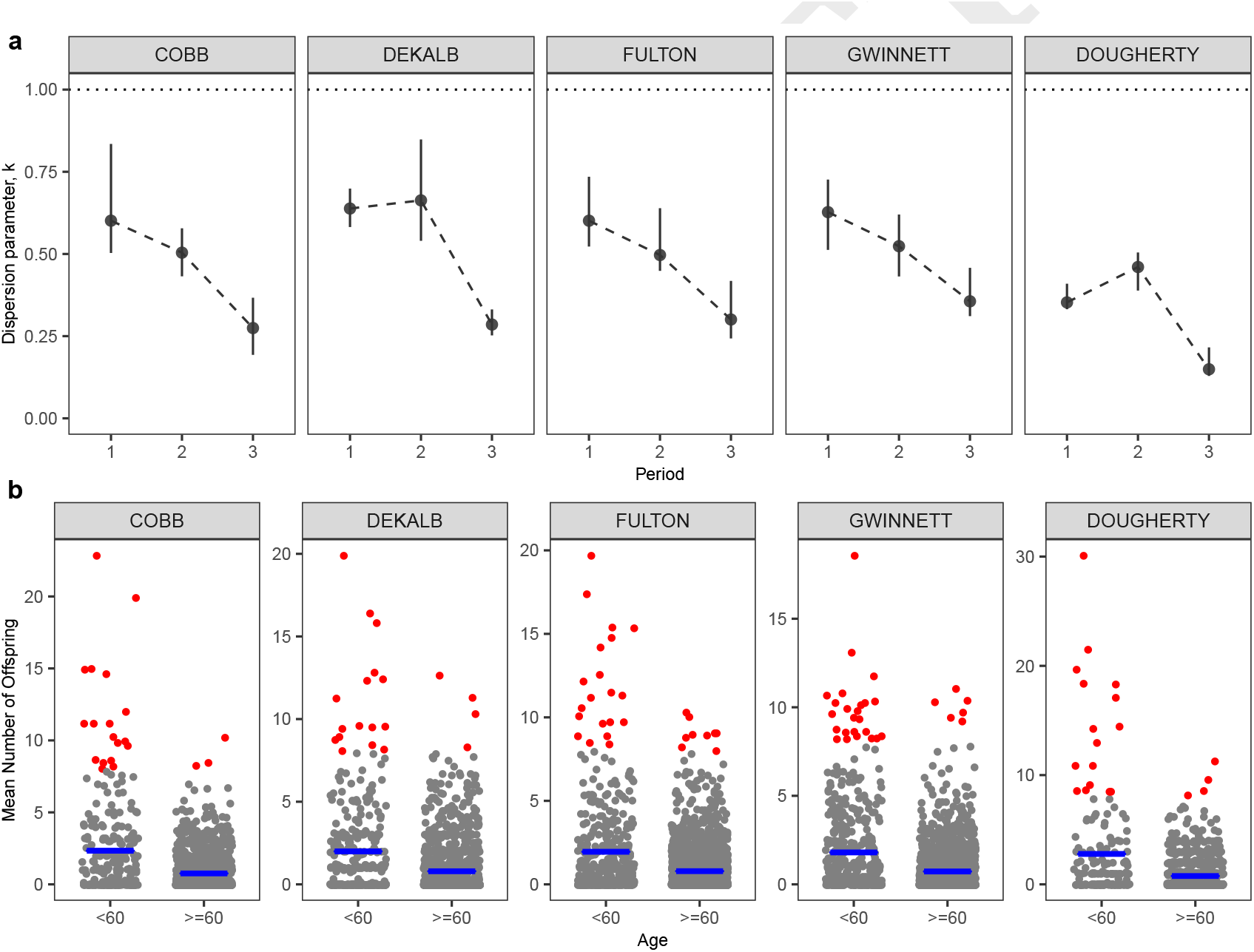
(a) Degree of super-spreading quantified by the dispersion parameter *k* (where *k* < 1 indicates significant super-spreading) during different periods of the outbreak. Let *T* be the day of announcing shelter-in-place order: period 1 is time *t* < *T*, period 2 is [*T*, *T* + 14) and finally period 3 is *t* > *T* +14. Overall, about top 2% of cases (that have highest mean number of offsprings) directly infected 20% of the total infections. (b) Mean number of offsprings generated by cases in each age group. Red dots represent those case have mean offspring >=8. The younger age group (<60) tends to have more cases that produce extreme number of offsprings, and also a larger average (blue lines) of the mean number of offsprings. Overall means of *k* also tend to be similar or smaller among the younger group: 0.53 for the younger group versus 0.82 for the older group in Cobb county, 0.57 versus 0.54 in DeKalb, 0.46 versus 0.64 in Fulton, 0.6 versus 0.6 in Gwinnett, and 0.39 versus 0.62 in Dougherty.

**Fig. 3.**
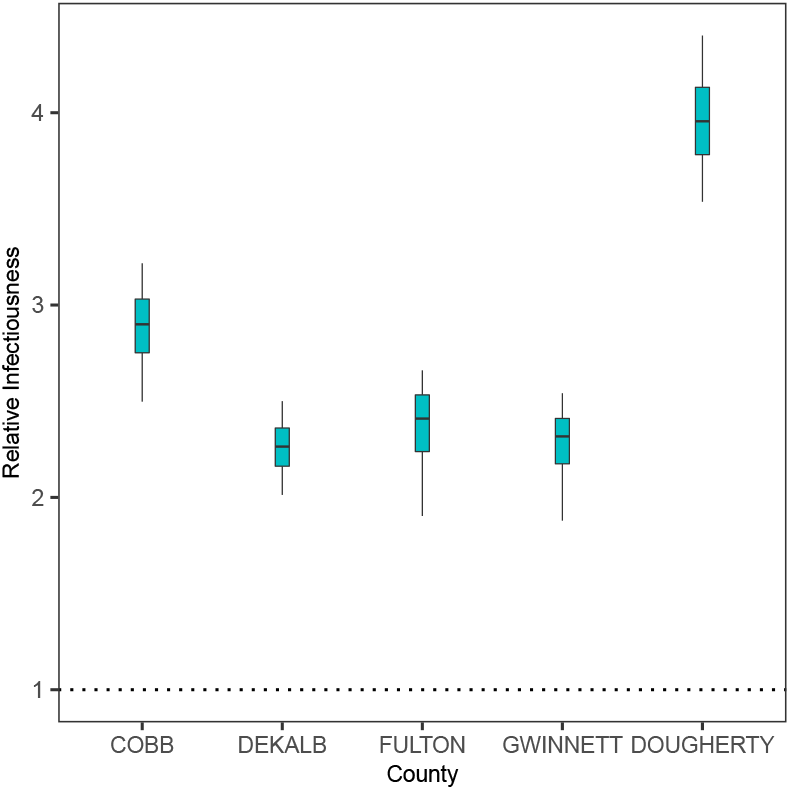
Infectiousness of younger patients (<60) relative to the older patients (>=60).

### Age-specific Infectiousness

A markedly low proportion of cases among the younger cases is observed in the current COVID-19 pandemic, indicating heterogeneity of susceptibility among different age groups [3, 19, 20]. Much is unknown about the variation of infectiousness among different age groups [1–3]. Our results suggest that the younger cases (<60) may be overall 2.78 [2.10, 4.22] times more infectious than elderly cases (>=60). Due to the very small number of reported cases in children (e.g., <15), we do not consider a finer age stratification (see also *Discussion*). We also test the robustness of these results towards to under-reporting and take into account the discrepancy in the reporting rates of different age groups (see next section *Sensitivity Analysis*). We also test the robustness of our results towards assumptions concerning relative susceptibility of the younger age group (see *Materials and Methods* and *Sensitivity Analysis*). It is worth noting that we are not explicitly modelling biological factors such as viral loads that may potentially affect infectiousness. Instead, our model captures how likely a case may generate secondary cases (see *Materials and Methods*), which collectively and implicitly accounts for multiple factors including viral loads and contact rates.

### Sensitivity Analysis

Under-reporting is ubiquitous feature of epidemiological data, and is particularly so for COVID-19 due to, in particular, a substantial number of asymptomatic cases and the lack of testings at the earlier stages of the pandemic. In particular, older people may tend to be more susceptible and develop severe symptoms, and hence have a higher probability of being reported [3, 19, 20]. Such a discrepancy in reporting rates may potentially affect our estimation of age-specific infectiousness. We explore the effect of such under-reporting on our results under these probable scenarios: we assume in March probabilities of being reported for younger case (<60) and older cases are respectively 0.1 and 0.2; and to account for increased testing capacity, these probabilities in April increase to 0.3 and 0.6 respectively. Details of how to include under-reported cases are given in *Materials and Methods*. Figure 4 shows that the younger age group remains to be more infectious than the older age group. The estimated impact of super-spreading also appears to be robust: estimated overall dispersion parameter *k* is 0.45 for Cobb county, 0.43 for Dekalb, 0.39 for Fulton, 0.49 for Gwinnett, and 0.32 for Dougherty.

**Fig. 4.**
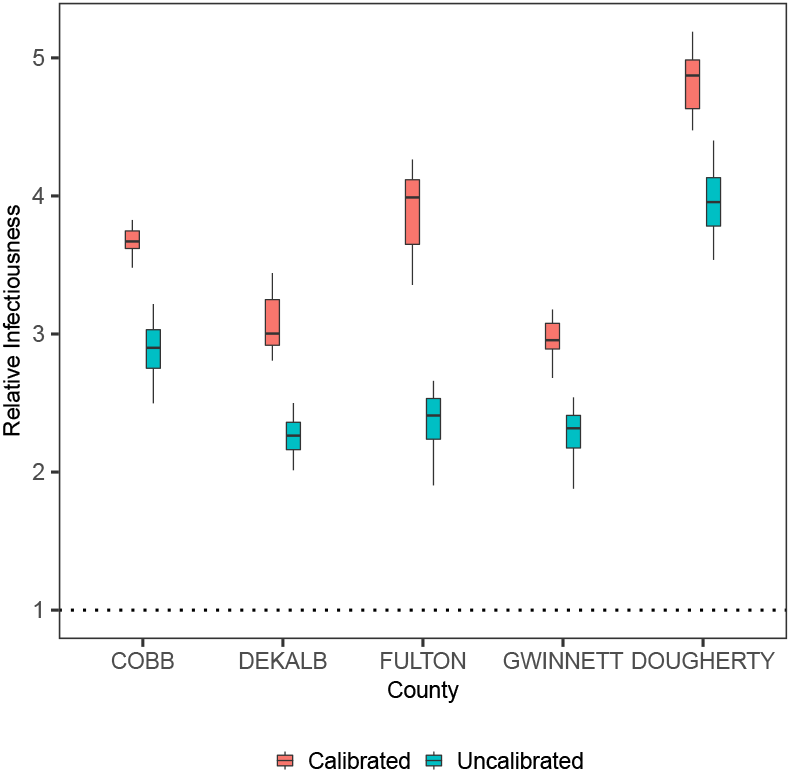
Infectiousness of younger patients (<60) relative to the older patients (>=60), calibrated for under-reported cases.

We assumed that the older age group is twice as susceptible as the younger group (see *Materials and Methods*). To explore if this assumption has an effect on the estimate of the relative infectiousness of the younger group, we fit the model by assuming equal susceptibility between the two age groups. The estimated infectiousness of the younger group relative to the older group is 2.84 [1.65, 3.60] which is similar to the estimate when we assume non-uniform susceptibility.

## Discussion

Transmission dynamics of infectious diseases are often non-linear and heterogeneous over space and time. It is important to exploit available data that are relevant to describing and estimating such complex processes. For COVID-19, a key step is to enable individual-level model inference that is able to statistically synthesize these datastreams, beyond aggregate-level dynamics [2, 11, 12]. In this paper, we incorporated multiple valuable datastreams including formal surveillance data into our individual-level spatio-temporal transmission modelling framework, achieving a more granular mechanistic understanding of the dynamics and heterogeneities in the transmission of SARS-CoV-2.

Our results give similar estimates of important population-level epidemiological parameters such *R*_0_ found in the literature, and reinforce the conclusion from most studies that social distancing measures are effective. This paper also advances our understanding of individual-level heterogeneities in the transmission of SARS-CoV-2, which is crucial for informing optimal interventions. We show that super-spreading is an important and ubiquitous feature throughout the pandemic, and it may have a pivotal role in driving large outbreaks outbreak in rural areas. The increasing significance of super-spreading over time also highlights the importance of maintaining social distancing measures that may curtail close contacts (e.g. gatherings with densely packed crowds). We also find that infected younger cases (<60) tend to be more infectious and to promote super-spreading. Our results have important implications for designing more effective control measures – particularly, highlight the importance of more targeted interventions.

Our study has a number of limitations. First of all, due to the lack of widely available testings, the under-reporting rate was almost surely high during earlier phases of the pandemic. Also, severity of symptoms (and hence reporting rates) may vary among different age groups. We explore the robustness of our main results towards these possible under-reporting scenarios in the *Sensitivity Analysis*. Reassuringly, our main conclusions appears to be largely robust. Second, with the lack of detailed individual movement data, our model implicitly assumes transmission occurred mostly near people’s homes. Nevertheless, most activities (hence transmissions) were likely to have clustered around homes due to the increasing public concern over the pandemic and announcements of shelter-in-place order back in March and April. For example, Fulton county, one of the counties with the highest number of cases, closed its school as early as March 10, about one month before the shelter-in-place order on April 3. As activities outside homes (e.g. gatherings at pubs) may tend to facilitate clusters of transmissions and super-spreading, our model may have thus underestimated both the infectiousness of younger adults who are more socially active and the degree of super-spreading. Third, we only consider modelling age-specific infectiousness in two age groups (<60 and >=60). The model would tend to be over-parameterized if we further break down the age groups that we have considered (e.g. by having a group for younger than 15), mainly due to imbalance of reported number of cases between age groups (particularly, markedly low reported numbers in very young population). And given the very few cases among the younger children, our results are mostly relevant for younger adults and elderly. Our current binning of age is still useful as the first group tends to be more socially active and is useful for informing the design of social distancing measures. Finally, although our analysis reveals the importance of age as a demographic risk factor of super-spreading, future work in linking them directly with biological factors (e.g., age-specific viral loads) may shed further light.

## Materials and Methods

### Spatio-temporal Transmission Process Model

We formulate an age-specific spatio-temporal transmission modelling framework that allows us to infer the unobserved infection times and transmission tree among cases, integrating detailed individual-level surveillance data, geo-spatial location data and highly-resolved population density (grid-)data, and aggregate mobility data. This approach also allows us to infer explicitly the distribution of the *offspring* (i.e. number of secondary cases generated) of each case. Our framework represents an extension of the models we developed *et al*. [8, 9], which were validated generally and applied successfully to dissect the transmission dynamics of the Ebola outbreak in Western Africa between 2014-2016. Figure 5 gives a schematic overview of the model.

**Fig. 5.**
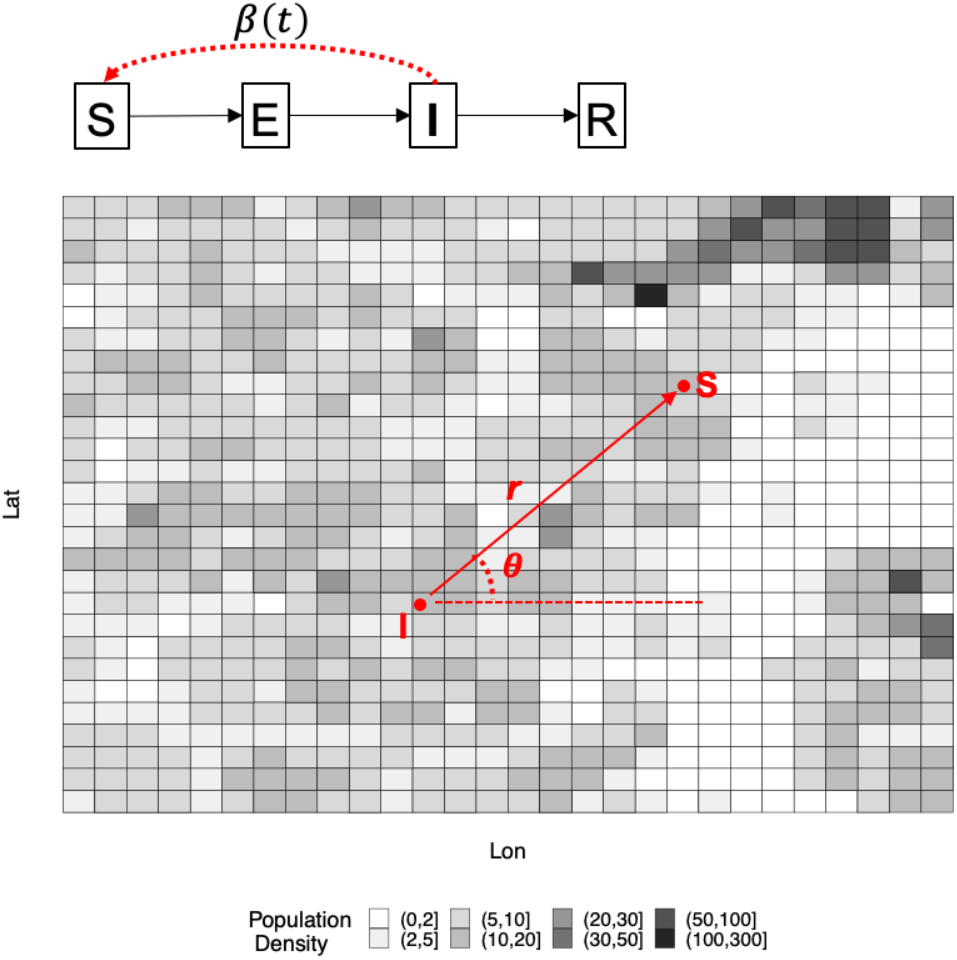
Schematic description of our model. We model individual-level transmission of SARS-CoV-2, in continuous time and space and over a heterogeneous landscape with varying population density over 100 × 100*m* grids. Disease status of an individual is assumed to follow the Susceptible-Exposed-Infectious-Recovered framework. Infectiousness of an infectious individual *β*(*t*) is time-dependent and decreases due to social distancing. Likelihood of transmission from the infectious individual to a susceptible individual, at distance *r* and angle *θ* measured from the infectious source, is determined by (1) a spatial movement kernel (density) function *f* with mean *η*, (2) change of the mean movement distance due to shelter-in-place order (informed by the aggregate mobility data), and (3) spatial distribution of population denoted by *ŝ* (i.e. detailed grid-level population density shown in the figure), which all together could more realistically account for heterogeneous mixing of individuals in the population [9].

More specifically, we model the occurrence of a new infection occurs as the first event in a non-homogeneous Poisson process with a time-varying rate *r*(*t*) = *n*(*t*) × *β*(*t*), where *n(t)* is number of infectious individuals at time *t* and *β*(*t*) the time-dependent infectiousness of a case. We consider that *β*(*t*) remained constant (i.e. the baseline infectiousness) before the time of state-wide shelter-in-place order was announced, and declined exponentially after the order according to a rate *ω*. We also allow a primary infection rate *α* which may account for infections that are not explicitly modelled (e.g. noise or imported cases). We further assume that the two age groups (<60 and >=60) have their own (free) baseline infectiousness parameters to allow for age-specific infectiousness.

Spatially, it is assumed that the probability of the new infection being at a certain position (polar coordinates measured by distance *r* and direction *θ*) away from the source of infection, is determined by the movement patterns of infectious individuals and the population density. Specifically, *r* and *θ* are drawn from an appropriate density function *g(r,0; n*, ***s***) (Ref. [9]). Details of *g*(*r*, *θ*; *η*, *ŝ*) are also given in the *SI Appendix, SI Text*. Recalled that *ŝ* is the population density (within many 100 × 100meter grids) across a county and *η* is the mean of the spatial kernel *f*. The mean of movement distance *η*, after the issuing of the shelter-in-place order, is assumed to change according to the county-wise percentage change of movement distance (which is computed from the Facebook mobility data, see below). We calculated the average distance of all trips per day occurring in a county from March 23 to April 2, 2020, to establish baseline mobility in distance prior to implementation of state-wide social distancing measures. We compared baseline mobility with mobility during the period from April 3 to May 5, after implementation of social distancing measures. Mobility data from Fulton, DeKalb, Gwinnett, and Cobb counties was readily available; data from Dougherty county was not. As a proxy for mobility in Dougherty county, we used data from Liberty and Glynn counties, which are also classified as ‘Small Metro’ according to the 2010 U.S. Census Bureau urbanicity classifications [21].

A new infection would go through an Exponentially-distributed incubation period with a mean parameter i, before showing symptoms and becoming infectious. It is assumed that patients <60 and >=60 have respectively probabilities 0.06 and 0.17 to be hospitalized [22]. The older age group is also assumed to be twice as susceptible as the younger group (<60) [3]. The sojourn time between symptom onset to hospitalization follows *Exp*(*c*). A non-hospitalized case is assumed to follow an Exponential distribution with a mean 14 days before recovery.

We estimate **Θ** (i.e. the parameter vector) in the Bayesian framework by sampling it from the posterior distribution *P*(**Θ** |**z**) where **z** are the data [8, 9, 23, 24]. Denoting the likelihood by *L*(***θ***; **z**), the posterior distribution of **Θ** is *P*(**Θ**|**z**) (X *L(****©***; ***z****)n(****©***), where n(**Θ**) is prior distribution for **Θ**. Non-informative uniform priors for parameters in **Θ** are used (see *SI Appendix*). Markov chain Monte Carlo (MCMC) techniques are used to obtain the posterior distribution. The unobserved infection times and transmission network and missing symptom onset dates are also imputed in the MCMC procedure. Details of the inferential algorithm are referred to *SI Appendix, SI Text*. Posterior distributions of parameters are given in *SI Appendix, SI Tables*.

### Sensitivity analysis

The number of total under-reported cases *m* for a particular age group during a particular period is calculated as *m = n/p* − *n*, where *n* is the reported number of cases and *p* the probability of being reported. We consider these probable scenarios: in March probabilities of being reported for younger case (<60) and older cases are respectively 0.1 and 0.2; and to account for increased testing capacity, these probabilities in April increase to 0.3 and 0.6 respectively. The *m* cases are then assigned infection times and spatial locations according to the temporal and spatial distributions of observed cases in the time period, before merging with the observed data. The infection times and sources of infections of the *m* cases are also treated as unknown and and are inferred in the data-augmentation procedure. Our main focus is to test how the potential discrepancy in reporting rate between age groups may impact our estimation of age-specific infectiousness.

## Data Availability

The authors do not own the surveillance data and cannot make it freely available. Data enquiry should be directed to the Georgia Department of Public Health.

## ACKNOWLEDGMENTS

We thank Dr. Laura Edison from the Georgia Department of Public Health for her efforts in leading and coordinating the research partnership between GDPH and Emory University. We also would like to acknowledge Nishant Kishore and the COVID-19 Mobility Network for their support accessing and analyzing Facebook mobility data. We also thank Professor Gavin Gibson for his helpful discussion during the preparation of this manuscript. Finally, we thank Google-For-Education program (*https://edu.google.com*) for donating free cloud credits which are used for geocoding involved in the research. BAL and KNN acknowledge support from NIH/NIGMS (R01 GM124280 and 3R01GM124280-03S1) NIH/NIAID (3R01AI143875-02S1) and NSF (2032084)

